# Functional plasticity of the olfactory network following functional septorhinoplasty in persistent COVID-19-related olfactory dysfunction: an fMRI study

**DOI:** 10.64898/2026.09.09.26362623

**Authors:** Aagat Sharma Khatiwada, Laura Mancini, Alfonso Luca Pendolino, Claudia A. M. Gandini Wheeler-Kingshott, Peter J Andrews

## Abstract

**Background:** Persistent COVID-19-related olfactory dysfunction (C19OD) has a significant disease burden. The underlying pathology is unclear, both at the level of the olfactory epithelium, and the higher neural networks. Functional septorhinoplasty (fSRP) is a novel intervention aimed at restoring odorant airflow to the olfactory epithelium, demonstrating clinically meaningful olfactory improvements in persistent C19OD patients. In the same cohort, we investigated the functional plasticity within the primary and higher-order olfactory brain networks using task-based functional magnetic resonance imaging (fMRI) before and after fSRP.

**Methods:** In a prospective cohort study, patients with persistent C19OD underwent psychophysical olfactory testing, nasal airflow measurements, and olfactory task-based fMRI at baseline and 6 months post-fSRP. Whole-brain fMRI data was analysed to evaluate changes in neural activation and their correlations with clinical parameters. Results were mapped to standardized anatomical and olfactory atlases.

**Findings:** Eight participants completed pre- and post-operative fMRI assessments. At baseline, increased deactivation was seen in the left hippocampus, and increased activation in bilateral cerebellum. Task-based activation was positively correlated with nasal volume in the right thalamus, and left cerebellum lobule VI. Post-operatively, task-based deactivation was seen in the left cerebellum crus II. Reduced task-dependent activation post-compared to pre-operatively was seen in the bilateral insula. Increased activation post-compared to pre-operatively positively correlated with change in nasal airflow in the right orbitofrontal cortex (OFC), superior and middle frontal gyrus, and frontal pole (FP). Increased post-compared to pre-operative activation correlated negatively with change in olfactory scores in bilateral insula, and left FP.

**Interpretation:** Persistent C19OD is characterized by widespread functional alterations across higher-order cortical regions, reflecting altered central olfactory processing. fSRP elicits significant neural plasticity within these higher-order olfactory areas. The findings support the mechanism that augmenting airflow to the olfactory cleft drives olfactory recovery through a combination of bottom-up epithelial input restoration and top-down network modulation.

## INTRODUCTION

Olfactory dysfunction (OD) is a common and debilitating sequela of SARS-CoV-2 (COVID-19) affecting up to 43% of infected cases since the pandemic.^1^ While spontaneous recovery occurs in the majority of cases, 8% of individuals experience persistent COVID-19 related OD (C19OD) lasting more than 3 years.^2^ This accounts for a significant disease burden with a considerable impact on quality of life.^3^

So far, the causes of C19OD remain unknown although some potential mechanisms include, peripheral destruction of the olfactory epithelium (OE) with reduced activation of olfactory sensory neurons (OSNs), degeneration of central olfactory pathways or a combination of both.^4–6^

The neural basis of olfactory processing involves a network of primary and higher-order cortical regions.^7^ OSNs first synapse in the olfactory bulb (OB), a paired structure sitting dorsal to the cribriform plate. From the OB, the axons of second order neurones, the mitral and tufted cells, extend to targets within the brain.^7^ The piriform cortex, amygdala, and entorhinal cortex receive direct neuronal stimulation from the OB, and form the primary olfactory cortex.^7^ These areas relay information to the secondary olfactory cortex, which includes the orbitofrontal cortex (OFC), insula, hippocampus, thalamus, hypothalamus, ventral striatum and pallidum.^8^ Other cortical regions including the superior frontal gyrus (SFG), middle frontal gyrus (MFG), and frontal pole (FP) also play an important role, mediating higher-order functions related to olfaction.^9^

Functional and structural plasticity within the human olfactory network has been demonstrated in response to sinonasal treatments targeting OD of various aetiologies which restore the natural function of the olfactory epithelium.^10,11^ Surgical treatment for chronic rhinosinusitis (CRS) and non-CRS related OD improves olfactory epithelial function. The mechanism of action is through treating and reducing the inflammatory burden at the nasal epithelial level including the olfactory epithelium which increases upstream olfactory functional activity within the insula, OFC, and other cortical areas, accompanied by changes in grey matter (GM) volume.^10,11^

Functional septorhinoplasty (fSRP) is a routine NHS surgical procedure performed to correct a structural obstruction causing nasal blockage. The two common causes of structural nasal obstruction include a deviated nasal septum (DNS) and internal nasal valve (INV) insufficiency. The septum is the midline structure which divides the 2 nostrils and the INV represents the narrowest area within the nose which helps regulate olfactory nasal airflow.^12^ fSRP typically increases nasal airflow by correcting the DNS and importantly augmenting or widening the INV through the use of spreader grafts.

Functional changes detected using resting state functional magnetic resonance imaging (fMRI) have been reported in people with persistent C19OD compared to people who spontaneously recovered olfaction and healthy controls.^13^ However, neural changes at brain network level, underpinning psychophysical improvements following treatment, have not previously been reported.

In a cohort of patients with persistent C19OD, we previously showed that fSRP resulted in improvement of olfactory function through increasing odorant delivery to the olfactory epithelium which increases activity dependent functionality of the OSNs.^14^ In this prospective controlled study, 67% of patients showed improvement above the minimal clinically important difference (MCID). In the same cohort, we investigated functional plasticity of the olfactory network using task-based fMRI before and after fSRP. We report the findings in this present study.

## MATERIALS AND METHODS

This fMRI analysis forms part of a larger prospective cohort study in patients with persistent C19OD undergoing fSRP.^14^ Participants were recruited from the long-COVID smell clinic at the University College London Hospitals, London, United Kingdom, between October 2022 and May 2023. This study received NHS ethical approval (REC ref 14/SC/1180) and was conducted in accordance with the Declaration of Helsinki. All participants provided full informed written consent prior to participation.

The inclusion and exclusion criteria for participation are listed in Table 1. Those meeting the eligibility criteria were offered fSRP, and were assessed at timepoints including baseline (T0), and 6-months (T2).

**Table 1.** Study inclusion and exclusion criteria.

| **Inclusion** | **Exclusion** |
| --- | --- |
| Age $\geq 18$ | Presence of other causes leading/contributing to OD (also confirmed by MRI of the head/sinuses) <sup>a</sup> |
| Aetiology of OD following a polymerase chain reaction—confirmed diagnosis of SARS-CoV-2 infection | History of PIOD prior to COVID-19 |
| OD confirmed at Sniffin' Sticks and longer than 18 months | Prior nasal/sinonasal/skull base surgery |
| OD failing to improve on conservative treatments, including OT and oral/topical corticosteroids | Bleeding disorders |
| Aesthetically unacceptable nasal deformity or reduced nasal airflow caused by a confirmed DNS and/or INV/ENV dysfunction | Blood thinners assumption |

**Abbreviations:** DNS, deviated nasal septum; ENV, external nasal valve; INV, internal nasal valve; MRI, magnetic resonance imaging; OD, olfactory dysfunction; OT, olfactory training; PIOD, post-infectious olfactory dysfunction.
Note: <sup>a</sup> These include: congenital olfactory loss, post-traumatic olfactory dysfunction, chronic rhinosinusitis, neoplasms, previous chemotherapy or radiotherapy to the head and neck, neurodegenerative diseases.

### Psychophysical testing and nasal measurements

All participants underwent clinical assessment, and psychophysical testing at T0 before fSRP, and again at T2. The median interval between T0 and the intervention was 55 days. Nasal airflow was measured bilaterally and unilaterally using peak nasal inspiratory flow (PNIF), obtained pre- and post-decongestion to reduce any influence of the nasal cycle.^15^ Similarly, acoustic rhinometry (AR) was used to obtain right and left nasal volume (NV), from which a total NV was calculated.

Psychophysical olfactory testing was performed using the Sniffin’ Sticks (S’S) tool [a measure of composite (TDI) and individual odour threshold (T), discrimination (D) and identification (I).^16^ Olfactory testing was performed birhinally. Normosmia was attributed where TDI was ≥30.75, hyposmia where TDI is >16, but <30.75, and functional anosmia ≤16.^17^ The minimal clinically important difference (MCID) for T, D and I are ≥2.5 points, ≥3 points and ≥3 points respectively, and ≥5.5 points for composite TDI.^18^

### Functional septorhinoplasty

All patients underwent fSRP using a standardised external approach, involving septoplasty with nasal bone realignment, internal nasal valve augmentation using spreader grafts (autologous cartilage), and columellar strut (autologous cartilage).

### Functional MRI

All participants in the intervention arm underwent fMRI at T0 and T2. As handedness does not appear to affect passive olfactory processing,^19^ patients were not excluded based on handedness alone. Patients not available for follow-up testing were excluded from the study. All subjects were asked to refrain from smoking, eating or drinking (except water) for 1 h prior to their assessment session, consistent with previously published olfactory-imaging protocols.^11^

### Imaging acquisition

Whole brain MRI was performed using a 3-T scanner (MAGNETOM Prisma, Siemens, Erlangen, Germany) with 64-channel head coil.

All participants underwent olfactory fMRI, as follows:

1. The fMRI protocol was previously described by Whitcroft et al. 2023.^20^ Two odorants were used for the fMRI (one per functional run): banana (odour 1) and cut grass (odour 2). Odour was delivered birhinally. Each participant underwent two functional runs per scanning session, with order of first odour pseudorandomised and counter-balanced across participants. At the end of each functional run, participants were asked to rate odour intensity (0–10, 10 = strongest) and hedonic valence (−5 to +5, +5 = most pleasant). Functional data were collected using a 2D GE-EPI sequence repetition time (TR) 1,550 ms, echo time (TE) 26 ms, field of view (FOV) 200 mm, flip angle (FA) 75°, voxel size 2.5 mm× 2.5 mm× 2.5 mm (in total, 50 slices).
2. Sagittal T1-weighted images were acquired using a 3-dimensional magnetization-prepared rapid acquisition gradient echo (MPRAGE) sequence. The following parameters were used: TR, 2,000 ms; TE, 1.96 ms; inversion time (TI), 880 ms; FOV, 282 mm× 282 mm; matrix size, 256 × 256; one slab, 208 slices per slab; voxel size, 1.1 mm× 1.1 mm× 1.1 mm; and FA, 8°.

### Imaging analysis

Functional data was analysed using SPM25 (Wellcome Centre of Imaging Neuroscience, UCL, London, United Kingdom) and MATLAB (The MathWorks, Natick, MA, United States). Pre-processing involved initial realignment and unwarping of functional images followed by segmentation of T1-weighted images according to SPM tissue probability maps. Co-registration of functional to anatomical images was then performed, as well as normalisation of all images to MNI space. Finally, functional data were smoothed using an 8 mm FWHM kernel. The structural images were averaged to obtain a group T1 image of the patient cohort at both time points. A first level general linear model analysis was performed, including the head motion correction parameters in the model. The conditions “odour > baseline” for each odour in each visit, and “post-op > pre-op” for each odour were modelled for each subject, using the canonical haemodynamic response function. Resultant contrast images were then subjected to a second level random-effects analysis with one-sample T-tests. The conditions baseline > odour and pre-op > post-op were tested at the second level analysis with a negative contrast.

A voxel-wise regression of various covariates was performed in the one-sample T-test at the second level, testing the correlation between the specific covariate and contrasts from the first level analysis. The covariates considered were the demeaned peak nasal inspiratory flow (PNIF), nasal volume (NV), threshold discrimination identification (TDI), and perceived pleasantness of odours (pleasantness_O1_ or pleasantness_O2_). When comparing post-op > pre-op, the following covariates were considered: post-operative PNIF minus pre-operative PNIF (ΔPNIF), post-operative NV minus pre-operative NV (ΔNV) and post-operative TDI minus pre-operative TDI (ΔTDI). For the regression analysis, the covariates being tested were included as regressors, along with the mean value of the effect being tested.

Conceptually, some covariates are related. For example, TDI is a more complex measure of perceived odour intensity and for this reason only TDI was included as a covariate. PNIF is physiologically dependent on to the NV. Collinearity between the covariates was tested with the Pearson’s product-moment correlation (IBM SPSS Statistics for Windows 10 (IBM Corp., Armonk, N.Y., USA), which measures the linear association between each pair of covariates. For each covariate the variance inflation factor (VIF) was calculated with the following methodology. Each variable was centred (subtracting its mean), followed by a regress on all the other covariates in the set, using ordinary least squares (OLS) with an intercept. The R² of that regression was computed (measuring how much of the covariate’s variance is explained by the other covariates combined), and VIF calculated with the formula VIF = 1 / (1 - R²). A VIF of 5 means R² = 0.80 (80% of that covariate’s variance is explained by the others), and a VIF of 10 means R² = 0.90 (90% of that covariate’s variance is explained by the others). Covariates were included in the second level model only if the Pearson’s correlation was less than 0.6 and the VIF was less than 4.

All whole brain analyses were tested with correction for multiple comparisons at the exploratory threshold of uncorrected p_unc_ < 0.001 with a minimum cluster extent of k ≥ 10 voxels. From the resulting clusters, unless otherwise specified, those that had cluster level p-values of either p_FWE_ < 0.05 or p_unc_ <0.05 were considered significant. Exact p-values for each cluster have been reported in the respective tables. Cluster location was identified with comparison to the Harvard-Oxford Cortical and Subcortical Structural Atlas in MNI space. Reported clusters were also identified in the olfactory atlas from Gaviraghi et al.^21^

## RESULTS

Twelve patients underwent fSRP for persistent C19OD. Of these, ten participants completed fMRI assessment at T0 and 8 completed a post-operative fMRI assessment at T2, and were eligible for analysis in the present study. Baseline demographic and clinical data are summarised in Table 2. Clinical measurements of participants, including Sniffin’ Sticks (S’S) test results, PNIF, and NV, at baseline and 6-month follow-up are summarised in Table 3. More detailed description of the results, including clinical measures, and olfactory scores, have previously been reported.^22^

**Table 2.**
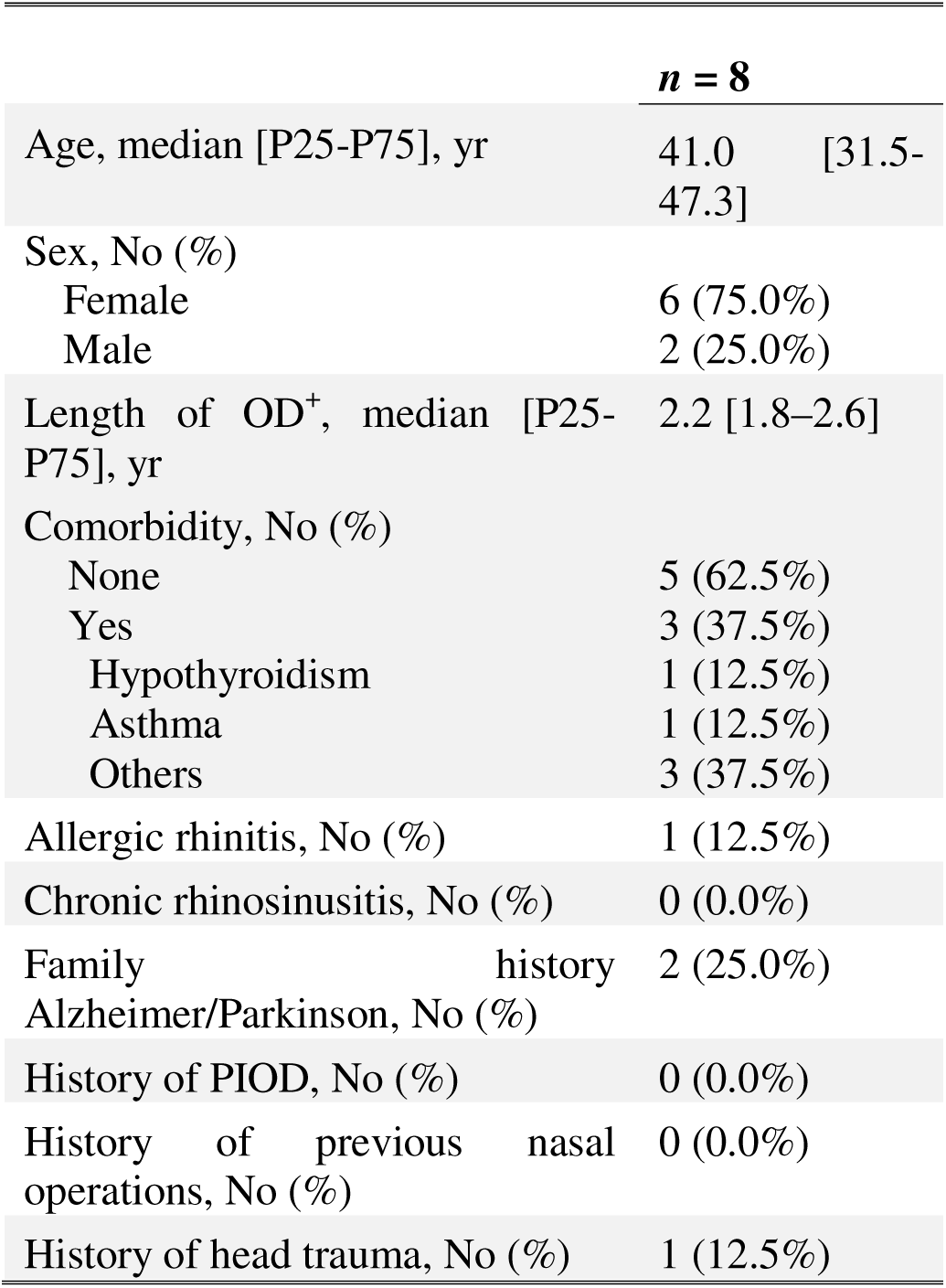
General characteristics of the population at baseline. +Length of OD is calculated as number of days from the infection date to the day of enrolment. OD: olfactory dysfunction; PIOD: post-infectious olfactory dysfunction.

**Table 3.** Clinical measurements of patients including Sniffin’ Sticks scores, peak nasal inspiratory flow (PNIF), and acoustic rhinometry (nasal volume, NV) at baseline and 6-month post-fSRP.

| n = 8 | Baseline (T0) | 6-month (T2) |
| --- | --- | --- |
| Sniffin' Sticks |  |  |
| TDI, median [P25-P75] | 22.3 [19.8–24.6] | 29.8 [24.4–30.6] |
| Threshold, median [P25-P75] | 2.3 [1.0–3.8] | 5.0 [3.6–6.5] |
| Discrimination, median [P25-P75] | 10.0 [9.8–10.5] | 12.0 [11.0–13.0] |
| Identification, median [P25-P75] | 9.5 [8.0–11.3] | 11.5 [9.8–13.0] |
| Normosmics, n (%) | 0 [0%] | 2 [25%] |
| Hyposmics, n (%) | 7 [87.5%] | 6 [75%] |
| Anosmics, n (%) | 1 [12.5%] | 0 [0%] |
| Nasal measurements |  |  |
| PNIF, median [P25-P75], L/min |  |  |
| Bilateral PNIF | 110.0 [87.5–133.8] | 170.0 [128.8–192.5] |
| Right PNIF | 62.5 [50.0–76.3] | 110.0 [72.5–126.3] |
| Left PNIF | 65.0 [56.3–77.5] | 77.5 [61.3–107.5] |
| Acoustic rhinometry, median [P25-P75] |  |  |
| Right Nasal volume, cm<sup>3</sup> | 5.9 [5.2–7.3] | 7.7 [5.2–9.8] |
| Left Nasal volume, cm<sup>3</sup> | 6.6 [6.0–7.8] | 8.1 [7.9–9.4] |

### fMRI Results – Odour 1

For odour 1, the variables that were not correlated with each other and that independently explained some variability of the fMRI data were: TDI and NV in the pre-operative MRI assessment; PNIF, TDI and NV in the post-operative MRI assessment; and ΔPNIF, ΔTDI, Δpleasantness in the pre-operative vs post-operative assessment.

#### Main effect of odour 1 inhalation and effect of covariates

Results for the main effect of odour 1 inhalation are summarised in Table 4. Pre-operatively, when considering areas with more activation when inhaling odour 1 than in the baseline condition, a cluster in the right cerebellum lobule VI reached significance (p_FWE_ = 0.004). Post-operatively, clusters were seen with more activation at baseline than when inhaling odour 1, in the right and left paracingulate gyrus (PCinG), right and left Heschl’s gyrus (HG), and in the left cerebellum (Crus II), along with four more clusters, two in the MFG and two in the SFG (Table 4).

**Table 4.**
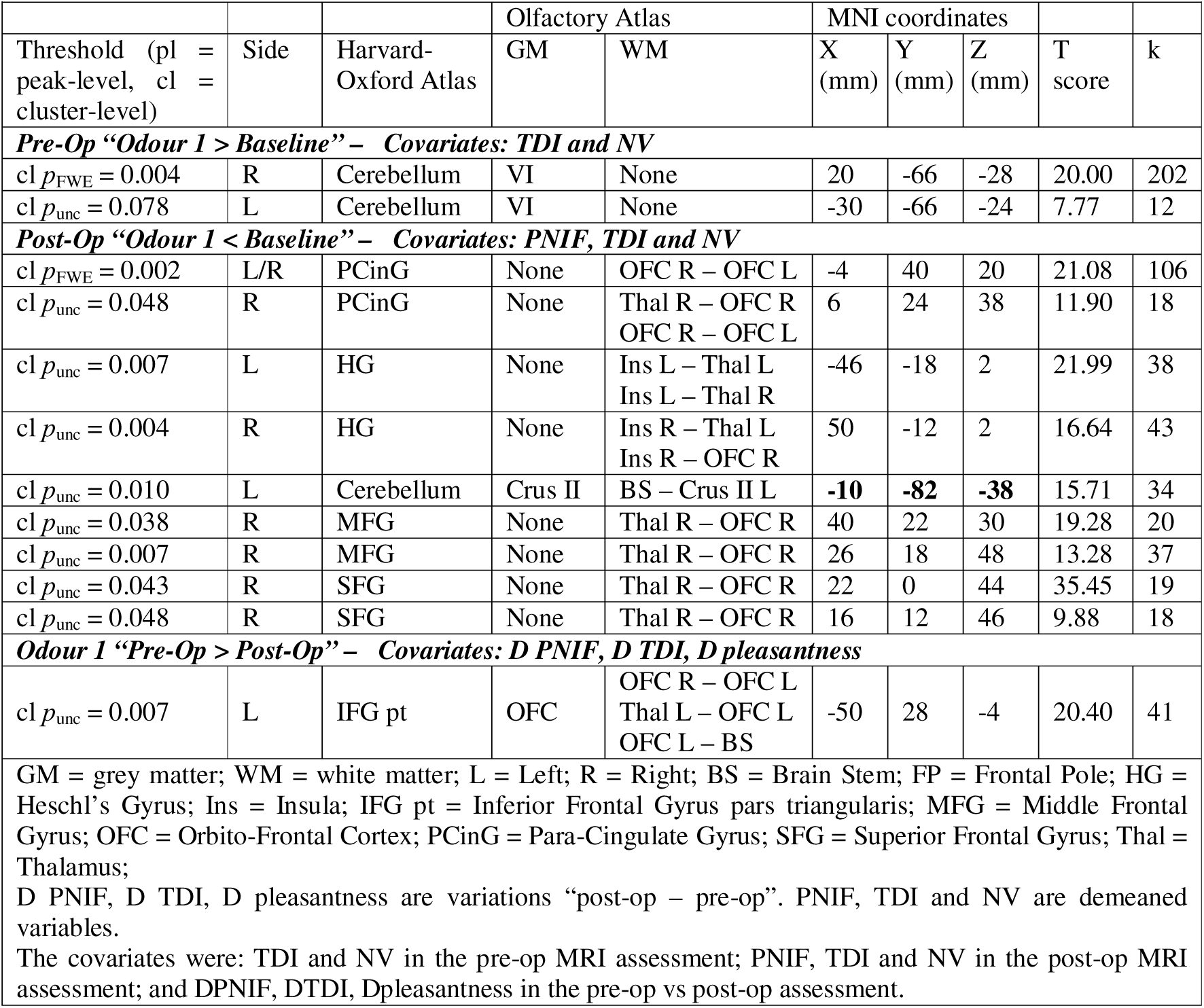
Odour 1 – Whole brain analysis – main effect – clusters that are also in the olfactory atlas.

When comparing pre-operative with post-operative activations, an area more active pre-operatively than post-operatively was observed in the left inferior frontal gyrus pars triangularis (IFG pt).

#### Voxel-wise regression with the covariates

Table 5 shows that pre-operatively there were three clusters where activation “Odour 1 > Baseline” was positively correlated with NV, located in the right cerebellum crus II, right thalamus and left superior parietal lobule (SPL).

**Table 5.** Odour 1 – Whole brain analysis – Voxel-wise regression with covariates – clusters also in the olfactory atlas.

|  |  |  |  |  |  |  |  |  |  |
| --- | --- | --- | --- | --- | --- | --- | --- | --- | --- |
| Threshold (pl = peak-level, cl = cluster level) | Side | Harvard-Oxford Atlas | Olfactory Atlas |  | MNI coordinates |  |  | T score | k |
|  |  |  | GM | WM | X (mm) | Y (mm) | Z (mm) |  |  |
| **Pre-Op – Covariate: NV, +ve contrast** |  |  |  |  |  |  |  |  |  |
| *Clusters where activation “Odour 1 > Baseline” was positively correlated uniquely with NV across subjects after eliminating the variance explained by TDI and by the mean effect of “Odour 1 > Baseline”* |  |  |  |  |  |  |  |  |  |
| cl $p_{FWE} = 0.016$ | R | Cerebellum | Crus II | BS – Crus II R | 20 | -66 | -28 | 33.34 | 153 |
| cl $p_{unc} = 0.028$ | R | Thal | Thal | Thal R – OFC R | 22 | -24 | 16 | 11.84 | 56 |
| cl $p_{unc} = 0.017$ | L | SPL | None | Hippo R – Thal L | -30 | -56 | 38 | 9.20 | 69 |
| **Post-Op – Covariates: TDI, -ve contrast** |  |  |  |  |  |  |  |  |  |
| *Clusters where activation “Odour 1 > Baseline” was negatively correlated uniquely with TDI across subjects after eliminating the variance explained by PNIF, NV and by the mean effect of “Odour 1 > Baseline”* |  |  |  |  |  |  |  |  |  |
| cl $p_{unc} = 0.034$ | L | Cerebellum | Crus I | BS – Crus I L | -34 | -76 | -30 | 20.54 | 21 |
| cl $p_{unc} = 0.038$ | R | Cerebellum | Crus II | BS – Crus II R | **10** | **-86** | **-32** | 11.38 | 20 |
| **Post-Op – Covariates: NV, -ve contrast** |  |  |  |  |  |  |  |  |  |
| *Clusters where activation “Odour 1 > Baseline” was negatively correlated uniquely with NV across subjects after eliminating the variance explained by PNIF, TDI and by the mean effect of “Odour 1 > Baseline”* |  |  |  |  |  |  |  |  |  |
| cl $p_{unc} = 0.001$ | L | Cerebellum | Crus I | BS – Crus I L | -34 | -76 | -30 | 30.26 | 60 |
| cl $p_{unc} = 0.015$ | L | Cerebellum | Crus II | BS – Crus II L | **-10** | **-82** | **-38** | 15.01 | 29 |
| **Post-Op > Pre-Op – Covariate: $\Delta$ PNIF +ve** |  |  |  |  |  |  |  |  |  |
| *Clusters where activation “post-op > Pre-Op” was positively correlated uniquely with $\Delta$ PNIF across subjects after eliminating the variance explained by $\Delta$ TDI, $\Delta$ pleasantness and by the mean effect of “post-op > Pre-Op”* |  |  |  |  |  |  |  |  |  |
| cl $p_{unc} = 0.003$ | R | FP | None | Thal R – OFC R<br>OFC R – OFC L | 22 | 58 | 22 | 21.69 | 52 |
| cl $p_{unc} = 0.038$ | R | FP | OFC | OFC R – OFC L | 36 | 60 | 2 | 17.88 | 22 |
| cl $p_{unc} = 0.035$ | R | FP | OFC | OFC R – OFC L<br>Thal R – OFC R | 44 | 42 | -2 | 11.04 | 11 |
| cl $p_{unc} = 0.038$ | R | SFG | None | Thal R – OFC R | 20 | 22 | 56 | 16.91 | 22 |
| cl $p_{unc} = 0.029$ | R | MFG | None | Thal R – OFC R | 44 | 22 | 26 | 16.90 | 25 |
| **Post-Op > Pre-Op – Covariate: $\Delta$ pleasantness +ve contrast** |  |  |  |  |  |  |  |  |  |
| *Clusters where activation “post-op > Pre-Op” was positively correlated uniquely with $\Delta$ pleasantness across subjects after eliminating the variance explained by $\Delta$ PNIF, $\Delta$ TDI and by the mean effect of “post-op > Pre-Op”* |  |  |  |  |  |  |  |  |  |
| cl $p_{unc} = 0.012$ | R | FP/PCinG | OFC | OFC R – OFC L | 4 | 58 | 6 | 14.81 | 35 |
| cl $p_{unc} = 0.052$ | L | FP/PCinG | OFC | OFC R – OFC L | -6 | 56 | 20 | 14.76 | 19 |
| GM = grey matter; WM = white matter; L = Left; R = Right; BS = Brainstem; EC = External capsule; FP = Frontal Pole; Hippo = Hippocampus; MFG = Middle Frontal Gyrus; OFC = Orbito-Frontal Cortex; PCinG = Para-Cingulate Gyrus; SFG = Superior Frontal Gyrus; SPL = Superior Parietal Lobule; Thal = Thalamus. D PNIF, D TDI, D pleasantness are variations “post-op – pre-op”. PNIF, TDI, NV are demeaned variables. The covariates were: TDI and NV in the pre-op MRI assessment; PNIF, TDI and NV in the post-op MRI assessment; and DPNIF, DTDI, Dpleasantness in the pre-op vs post-op assessment. |  |  |  |  |  |  |  |  |  |

Post-operatively, there were two clusters, one in the left cerebellum (crus I) and one in the right cerebellum (crus II), where activation “Odour 1 > Baseline” was negatively correlated with TDI. There were also two clusters in the left cerebellum, one in crus I and one in crus II, where activation “Odour 1 > Baseline” was negatively correlated with NV. Of these clusters, the one in the left cerebellum crus II with coordinates [-10, -82, -38] (in bold in tables 4 and 5) had as main effect the deactivation “Odour 1 < Baseline” and the residual variance negatively correlated with TDI and NV, indicating that worse clinical measures of lower TDI and lower NV had less deactivation as main effect.

For the contrast post-op > pre-op, five clusters, all in the right hemisphere, positively correlated with ΔPNIF across subjects: orbito-frontal cortex (OFC) (two clusters), SFG MFG, and FP. In two clusters, one in the right and one in the left para-cingulate gyrus portion of the OFC (the one on the left almost reaching significance with cluster level p_unc_ = 0.052) post-op > pre-op activation was positively correlated with Δpleasantness.

### fMRI Results – Odour 2

For odour 2, the variables that were not correlated with each other and that independently explained some variability of the fMRI data were: TDI and NV in the pre-operative MRI assessment; PNIF, TDI and NV in the post-operative MRI assessment; and ΔTDI, ΔNV and Δpleasantness in the pre-operative vs post-operative assessment.

#### Main effect of odour 2 inhalation and effect of covariates

Results for the main effect of odour 2 inhalation are summarised in Table 6. Pre-operatively, a cluster of activation in the left hippocampus was seen for odour 2 < baseline. Post-operatively, odour 2 < baseline activations were seen in the left and right precuneus cortex (PrCC), right OFC, right thalamus, right SFG, left parietal operculum cortex (POC), and left FP.

**Table 6.** Odour 2 – Whole brain analysis – main effect – only clusters also in the olfactory atlas.

|  |  |  |  |  |  |  |  |  |  |
| --- | --- | --- | --- | --- | --- | --- | --- | --- | --- |
| Threshold (pl = peak-level, cl = cluster-level) | Side | Harvard-Oxford Atlas | Olfactory Atlas |  | MNI coordinates |  |  | T score | k |
|  |  |  | GM | WM | X (mm) | Y (mm) | Z (mm) |  |  |
| **Pre-Op “Odour 2 < Baseline” – Covariates: TDI and NV** |  |  |  |  |  |  |  |  |  |
| cl $p_{unc} = 0.020$ | L | Hippo | Hippo | Thal L – Hippo L | -40 | -32 | -12 | 22.91 | 83 |
| **Post-Op “Odour 2 < Baseline” – Covariates: PNIF, TDI and NV** |  |  |  |  |  |  |  |  |  |
| cl $p_{FWE} = 0.039$ | L | PrCC | None | Thal R – Hippo L<br>Hippo L – Ins L<br>Hippo R – Thal L | -12 | -58 | 28 | 19.47 | 65 |
| cl $p_{unc} = 0.025$ | R | PrCC | None | Thal R – Hippo L<br>Hippo R – Ins R<br>Hippo R – Thal L | **12** | **-54** | **32** | 14.26 | 26 |
| cl $p_{FWE} = 0.030$ | R | OFC | OFC | OFC R – OFC L | 4 | 48 | 0 | 13.74 | 69 |
| cl $p_{unc} = 0.013$ | R | Thal | Thal | Thal R – OFC R | 22 | -22 | 14 | 62.40 | 33 |
| cl $p_{unc} = 0.016$ | R | SFG | None | Thal R – OFC R | 22 | 30 | 36 | 24.39 | 31 |
| cl $p_{unc} = 0.009$ | L | POC | None | Ins L – OFC L<br>Thal L – Ins L | -36 | -32 | 22 | 19.19 | 37 |
| cl $p_{unc} = 0.014$ | L | FP | None | OFC R – OFC L | -20 | 42 | 40 | 17.46 | 32 |
| **Odour 2 “Pre-Op > Post-Op” – Covariates: D TDI, D NV, D pleasantness** |  |  |  |  |  |  |  |  |  |
| pl $p_{unc} < 0.001$,<br>$\geq 10$ voxels | R | Ins | None | PirC R – Thal R<br>Thal R – OFC R | 40 | -4 | -10 | 15.49 | 19 |
| pl $p_{unc} < 0.001$,<br>$\geq 10$ voxels | L | Ins | None | Amy L – OFC L<br>PirC L – OFC L<br>Thal L – OFC L | -40 | 8 | -8 | 13.50 | 15 |
| pl $p_{unc} < 0.001$,<br>$\geq 10$ voxels | R | POC | None | Ins R – OFC R | 50 | -32 | 28 | 13.75 | 16 |
| GM = grey matter; WM = white matter; L = Left; R = Right; Amy = Amygdala; FP = Frontal Pole; Hippo = Hippocampus; Ins = Insula; OFC = Orbito-Frontal Cortex; PirC = Piriform Cortex; PrCC = Pre-Cuneus Cortex; POC = Parietal Operculum Cortex; SFG = Superior Frontal Gyrus; Thal = Thalamus. |  |  |  |  |  |  |  |  |  |
| D PNIF, D TDI, D pleasantness are variations “post-op – pre-op”. PNIF, TDI and NV are demeaned variables. |  |  |  |  |  |  |  |  |  |
| The covariates were: TDI and NV in the pre-op MRI assessment; PNIF, TDI and NV in the post-op MRI assessment; and DTDI, DNV, Dpleasantness in the pre-op vs post-op assessment. |  |  |  |  |  |  |  |  |  |

Comparing pre-operative with post-operative activations, only areas more active pre-operatively than post-operatively were observed, in bilateral insula, and right POC (with the peak level threshold of p_unc_ < 0.001 and cluster size ≥ 10 voxels)

#### Voxel-wise regression with the covariates

Table 7 shows that in the pre-operative session, a cluster was identified in the left cerebellum where odour 2 > baseline positively correlated with the NV.

**Table 7.** Odour 2 – Whole brain analysis – Voxel-wise regression with covariates – only clusters also in the olfactory atlas.

|  |  |  | Olfactory Atlas |  | MNI coordinates |  |  |  |  |
| --- | --- | --- | --- | --- | --- | --- | --- | --- | --- |
| Threshold (pl = peak-level, cl = cluster level) | Side | Harvard-Oxford Atlas | GM | WM | X (mm) | Y (mm) | Z (mm) | T score | k |
| **Pre-Op – Covariate: NV, +ve contrast** |  |  |  |  |  |  |  |  |  |
| Clusters where activation “Odour 2 > Baseline” was positively correlated uniquely with NV across subjects after eliminating the variance explained by TDI and by the mean effect of “Odour 2 > Baseline” |  |  |  |  |  |  |  |  |  |
| cl $p_{unc} = 0.030$ | L | Cerebellum | VI | BS – Cereb VI L | -18 | -68 | -22 | 8.50 | 71 |
| **Post-Op – Covariates: PNIF, -ve contrast** |  |  |  |  |  |  |  |  |  |
| Clusters where activation “Odour 2 > Baseline” was negatively correlated uniquely with PNIF across subjects after eliminating the variance explained by TDI, NV and by the mean effect of “Odour 2 > Baseline” |  |  |  |  |  |  |  |  |  |
| cl $p_{FWE} = 0.001$ | R | CinG pd | None | Thal R – OFC R | 12 | -22 | 46 | 43.64 | 127 |
| cl $p_{FWE} < 0.001$ | L | PrCC | None | Thal R – Hippo L | -8 | -68 | 34 | 32.53 | 140 |
| cl $p_{FWE} = 0.004$ | R | PrCC | None | Hippo R – Thal L | **12** | **-54** | **32** | 15.09 | 101 |
| cl $p_{unc} = 0.037$ | R | PCinG | None | OFC R – OFC L | 4 | 48 | 0 | 11.92 | 22 |
| cl $p_{unc} = 0.014$ | L | PCinG | None | OFC R – OFC L | -14 | 50 | 10 | 10.61 | 32 |
| **Post-Op – Covariates: TDI, -ve contrast** |  |  |  |  |  |  |  |  |  |
| Clusters where activation “Odour 2 > Baseline” was negatively correlated uniquely with TDI across subjects after eliminating the variance explained by PNIF, NV and by the mean effect of “Odour 2 > Baseline” |  |  |  |  |  |  |  |  |  |
| cl $p_{FWE} = 0.014$ | R | CinG pd | None | Thal R – OFC R | 12 | -22 | 46 | 29.33 | 81 |
| cl $p_{unc} = 0.045$ | L | CinG ad | None | OFC R – OFC L | -8 | 38 | 0 | 9.42 | 20 |
| cl $p_{FWE} = 0.021$ | L | PrCC | None | Thal R – Hippo L<br>Hippo R – Thal L | -8 | -68 | 34 | 24.05 | 74 |
| cl $p_{unc} = 0.016$ | R | PrCC | None | Hippo R – Thal L | **12** | **-54** | **34** | 20.64 | 31 |
| cl $p_{FWE} = 0.002$ | L | PCinG | None | OFC R – OFC L | -6 | 52 | 16 | 13.70 | 114 |
| cl $p_{unc} = 0.033$ | R | OFC | OFC | Thal R – OFC R | 42 | 28 | -6 | 14.82 | 23 |
| cl $p_{unc} = 0.025$ | L | FP | None | OFC R – OFC L | -24 | 34 | 14 | 13.61 | 26 |
| **Post-Op – Covariates: NV, -ve contrast** |  |  |  |  |  |  |  |  |  |
| Clusters where activation “Odour 2 > Baseline” was negatively correlated uniquely with NV across subjects after eliminating the variance explained by PNIF, TDI and by the mean effect of “Odour 2 > Baseline” |  |  |  |  |  |  |  |  |  |
| cl $p_{unc} = 0.030$ | L | Cerebellum | VI / Crus I | None | -18 | -78 | -22 | 12.58 | 24 |
| **Post-Op > Pre-Op – Covariate: $\Delta$ TDI -ve contrast** |  |  |  |  |  |  |  |  |  |
| Clusters where activation “Post-Op > Pre-Op” was negatively correlated uniquely with $\Delta$ TDI across subjects after eliminating the variance explained by $\Delta$ NV, $\Delta$ pleasantness and by the mean effect of “Post-Op > Pre-Op” | | | | | | | | | |
| cl $p_{unc} = 0.027$ | R | Ins | Ins | Thal R – OFC R | 40 | -4 | -10 | 20.79 | 43 |
| cl $p_{unc} = 0.012$ | L | Ins | None | Amy L – OFC L<br>Ins L – OFC L | -40 | 8 | -8 | 19.21 | 59 |
| cl $p_{unc} = 0.005$ | L | FP | OFC | OFC R – OFC L | -4 | 60 | 20 | 14.62 | 78 |
| cl $p_{unc} = 0.010$ | L | PrCC | None | Thal R – Ins L<br>Hippo R – Thal L | -10 | -54 | 46 | 12.42 | 62 |
| GM = grey matter; WM = white matter; L = Left; R = Right; BS = Brain Stem; Cereb VI = Cerebellum VI; CinG ad = Cingulate Gyrus, anterior division; CinG pd = Cingulate Gyrus, posterior division; FP = Frontal Pole; Hippo = Hippocampus; OFC = Orbito-Frontal Cortex; PrCC = Pre-Cuneus Cortex; PCinG = Para-Cingulate Gyrus; Thal = Thalamus. |  |  |  |  |  |  |  |  |  |
| D PNIF, D TDI, D pleasantness are variations “post-op – pre-op”. PNIF, TDI, NV are demeaned variables. The covariates were: TDI and NV in the pre-op MRI assessment; PNIF, TDI and NV in the post-op MRI assessment; and DTDI, DNV, Dpleasantness in the pre-op vs post-op assessment. |  |  |  |  |  |  |  |  |  |

In the post-operative session, clusters where activation for odour 2 > baseline negatively correlated with PNIF were identified in the right cingulate gyrus posterior division (CinG pd), bilateral PrCC, and bilateral PCinG. Similarly, clusters of activation negatively correlated with TDI in the right CinG pd, bilateral PrCC, left PCinG, right OFC, left FP, and left cingulate gyrus anterior division (CinG ad). A cluster of activation negatively correlated with NV was identified in the left cerebellum crus I/VI.

Of these clusters, the one in the right PrCC with coordinates [12, -54, 34], had as main effect (Table 6) odour 2 < baseline. In this cluster, the regression with covariates showed a negative correlation with PNIF and/or with TDI i.e. smaller deactivations correlated with the worse clinical measures of lower PNIF and/or lower TDI.

Post-Op > pre-Op activation negatively correlated uniquely with ΔTDI in bilateral insula, left FP, and left PrCC.

## DISCUSSION

To our knowledge, this is the first study to demonstrate functional plasticity within higher-order olfactory networks in patients with persistent C19OD following fSRP. We previous demonstrated that fSRP results in statistically significant, and clinically meaningful improvements in olfactory function in patients with persistent C19OD of over 2 years duration.^22^ The exact mechanism underlying this remains unclear. In the present study, fMRI findings demonstrate functional activation patterns at baseline in this cohort of patients. Additionally, we demonstrate changes in cortical activation following fSRP relative to pre-operatively, suggesting functional reorganisation and neuroplasticity within the higher-order olfactory network following the intervention.

### Long-standing olfactory circuit alterations in persistent C19OD

In our cohort of patients with persistent C19OD, we demonstrated functional alterations in cortical regions which have already been shown to be highly relevant in olfaction. At baseline, we observed increased activation as well as deactivation in response to odour inhalation in various brain areas. Increased deactivation, for example, was seen in the left hippocampus with odour 2. On the other hand, increased activation on odour 1 inhalation at baseline was seen in brain areas including the right cerebellum. Task-based activation to both odours was positively correlated with NV across subjects. For odour 1 this cluster was in the right thalamus, in close proximity to the fascicles connecting the right cerebellum with the brainstem, the right thalamus with the right OFC and the right hippocampus with the left thalamus. For odour 2 such cluster was in the left cerebellum lobule VI, in close proximity to the fascicle connecting the brainstem with the cerebellum lobule VI.

The OFC, hippocampus, and thalamus form part of a diverse cortical network involved in higher order olfactory processing, including olfactory working memory, emotional and hedonic processing of olfactory stimuli, semantic processing of odours, and multisensory integration of olfactory stimuli.^9,23,24^

Previous studies have reported both increases and decreases in functional connectivity (FC) of olfactory networks in persistent C19OD in these areas.^13^ For example, increased FC between the OFC and olfactory cortex, and the anterior insula and cerebellum, were seen in patients with persistent C19OD compared to controls. On the other hand, reduced FC was seen between the OFC and the dorsal anterior cingulate gyrus. Zhang et al.^25^ showed increased resting state connectivity within olfactory networks including the common nodes (SFG, MFG, and PrCC within the so-called default mode network, and amygdala and insula in the olfactory network) in persistent C19OD (over 3 months’ duration). Increased resting-state activity and connectivity in parts of the olfactory network might reflect compensation through increased reliance on higher-order processes such as olfactory recognition memory in persistent C19OD patients. Examples of top-down compensation for reduced olfactory function (specifically, poorer odour identification scores) through semantic memory have been seen in other olfactory areas.^26^

A systematic review looking at the resting state fMRI changes in C19OD noted considerable heterogeneity between studies, both in anatomical location and the direction of change, which could reflect a mechanism of disruption (and in some cases recovery) with a diverse timecourse..^27^ However, alterations were consistently seen in the broad cortical network underlying olfactory processing. The review also suggested that increased FC in areas including the OFC and piriform cortex might be a compensatory feature of early OD, noting that shorter durations of OD were associated with increased FC. Similarly, variation in functional responses in task-based fMRI studies of C19OD were also seen, although only 2 such studies were identified. One study found more robust trigeminosensory activity in C19OD compared to post-infectious OD (PIOD), whereas another found reduced activation in the upper frontal lobe and basal ganglia in C19OD compared to healthy controls.

These areas of functional change are consistent with changes seen in our own cohort of patients with C19OD lasting over 2 years. Gaviraghi et al., in their olfactory atlas, demonstrated the relevance of these areas in olfaction.^28^ Furthermore, through multisequence MRI, they demonstrated that those with persisting C19OD (mean duration of smell loss over 6 months), compared to healthy controls, showed neuroinflammation and axonal degeneration, and compared to those whose olfactory function recovered, myelin-related changes (such as myelin loss). These findings suggest direct pathological changes of the cortical olfactory network in persistent C19OD, and a potential mechanism suggested was autoimmune inflammation. Areas of the brain implicated in that study included the hindbrain (brainstem and cerebellum), and the secondary olfactory cortex, including the insula, OFC, hippocampus and thalamus. The functional changes reported in our present study are consistent with these brain areas.

Interestingly, in our present study, we also saw instances of increased task-dependent activation in persistent C19OD, including in several areas within the cerebellum (including bilateral cerebellum lobule VI, and right cerebellum crus II). These findings are especially pertinent in light of the findings of Gaviraghi et al. whereby there was a preponderance for alterations in the hindbrain, including brainstem and cerebellum, in those with persistent C19OD.^28^ While the role of the cerebellum in the olfactory pathway is unclear, it may be involved in alterations of sniffing behaviour and chemosensory integration.

### Changes in olfactory circuits in persistent C19OD following fSRP

Similar to widespread cortical changes seen at baseline in persistent C19OD, fSRP heralded significant plasticity in various brain areas, and included functional change in both directions, with increased activations and deactivations.

Reduced task-dependent activation was seen post-operatively compared to pre-op, in the left inferior frontal gyrus pars triangularis (IFG pt) (with odour 1), and bilateral insula (with odour 2). The IFG pt is in the left OFC (in the olfactory atlas) and close to the tracts connecting the right and left OFC, and the left OFC with the left thalamus and brainstem.

The insula is a central connecting hub in the olfactory pathway, and along with the OFC, and thalamus, is a key area involved in higher-order olfactory processing such as olfactory recognition memory.^29,30^ Reduced activation in these areas post-operatively may represent reduced reliance on these higher-order olfactory functions as a compensatory mechanism. Eek et al., for example, found functional suppression in brain areas including the insula during instances of successful odour recognition.^29^

Importantly, for odour 1, increased activation post-operatively compared to pre-operatively positively correlated with ΔPNIF in five clusters: right OFC, right SFG and MFG, and right FP, and in close proximity with fascicles connecting the left OFC with the right OFC and the right OFC with the right thalamus. On the other hand, clusters with increased post-operative compared to pre-operative activation with odour 2 correlated negatively with ΔTDI in bilateral insula, left FP, and left PrCC. These are all brain areas which have a recognised role in higher order olfactory processing, but the bidirectionality of the correlation demonstrates the complexity of the interactions within the olfactory neural network.

The neuroplasticity in response to fSRP demonstrated in the present study, and its correlation with change in nasal airflow, however, is consistent with previous reports, and involves some of the same brain areas previously recognised, such as the OFC and insula.^10^ Similarly, olfactory functional change resulting from functional sinus surgery for chronic rhinosinusitis (CRS) has also shown to be associated with plasticity in these higher cortical areas.^11^ CRS and persistent C19OD/PIOD represent different pathophysiological processes underpinning OD. Neural plasticity in similar cortical areas following surgical intervention for these respective conditions suggests that this plasticity is being mediated, at least in part, in a ‘bottom-up’ manner through increased stimulation of the peripheral olfactory apparatus.

Similarly, the bidirectionality of the functional changes, including both increased activations and deactivations at baseline and following fSRP, likely represents compensatory changes within a complex olfactory network, including top-down modulation.

For example, post-operatively, we showed task-based deactivation in the left cerebellum crus II, in close proximity to tracts connecting the cerebellum with the brainstem (BS). The same cluster also negatively correlated with TDI, and NV. Gaviraghi et al., in their cohort of persistent C19OD patients, demonstrated significant structural changes in the hindbrain (brainstem/cerebellum) networks, although the exact link between these structural changes and the functional alterations seen in our cohort remains to be elucidated.^28^ As suggested in that multimodal MRI study, changes in the olfactory network might represent direct pathology (e.g. neuroinflammation) mediated by long-term sequelae of COVID-19. It would be instructive to assess such structural changes in olfactory brain areas, including the brainstem-cerebellum network, in response to fSRP. This would provide clues towards the underlying mechanisms of change. While these exact mechanisms remain unclear, our findings provide evidence of the functional neuroplasticity mediated by fSRP in patients with persistent C19OD.

The principal limitation of this study was the small sample size. This reduces statistical power and limits the precision and stability of the estimated associations between post-operative changes in functional activity, PNIF and olfactory performance. The relatively large number of voxel-wise analyses and correlations also requires cautious interpretation. Although the reported associations were identified using appropriate statistical correction, small samples can produce unstable effect estimates, particularly when several odours, contrasts and clinical measures are examined. The findings should therefore be regarded as hypothesis-generating and require replication in larger cohorts.

## CONCLUSION

fSRP has previously been demonstrated to improve olfactory function in patients with persistent C19OD. In these patients with OD lasting over 2 years, a wide network of cortical areas including the OFC, thalamus, cerebellum, brainstem network, demonstrates functional alterations, in both increased and deactivated direction, revealing a complex mechanism of response to the injury caused by COVID-19. This suggests altered higher-order olfactory processing, and may be mediated by direct pathological processes within the olfactory network, or a combination of plasticity mediated due to changes at the level of the olfactory epithelium in a ‘bottom-up’ manner, and ‘top-down’ modulation. Following fSRP, odour inhalation elicited plasticity within higher-order olfactory regions. Importantly, areas of increased activation following fSRP correlated with increased nasal airflow, supporting the notion that increased nasal airflow to the olfactory cleft is responsible for some of the improvement in olfactory function.

## Data Availability

All data produced in the present study are available upon reasonable request to the authors

## REFERENCES

1. Saniasiaya J, Islam MA, Abdullah B. Prevalence of Olfactory Dysfunction in Coronavirus Disease 2019 (COVID-19): A Meta-analysis of 27,492 Patients. The Laryngoscope. 2021;131(4):865–878. doi:10.1002/lary.29286

2. Boscolo-Rizzo P, Spinato G, Hopkins C, et al. Evaluating long-term smell or taste dysfunction in mildly symptomatic COVID-19 patients: a 3-year follow-up study. Eur Arch Oto-Rhino-Laryngol Off J Eur Fed Oto-Rhino-Laryngol Soc EUFOS Affil Ger Soc Oto-Rhino-Laryngol - Head Neck Surg. 2023;280(12):5625–5630. doi:10.1007/s00405-023-08227-y

3. Saniasiaya J, Prepageran N. Impact of olfactory dysfunction on quality of life in coronavirus disease 2019 patients: a systematic review. J Laryngol Otol. 2021;135(11):947–952. doi:10.1017/S0022215121002279

4. Doty RL. Olfactory dysfunction in COVID-19: pathology and long-term implications for brain health. Trends Mol Med. 2022;28(9):781–794. doi:10.1016/j.molmed.2022.06.005

5. Vaira LA, Hopkins C, Sandison A, et al. Olfactory epithelium histopathological findings in long-term coronavirus disease 2019 related anosmia. J Laryngol Otol. 2020;134(12):1123–1127. doi:10.1017/S0022215120002455

6. Finlay JB, Brann DH, Abi Hachem R, et al. Persistent post-COVID-19 smell loss is associated with immune cell infiltration and altered gene expression in olfactory epithelium. Sci Transl Med. 2022;14(676):eadd0484. doi:10.1126/scitranslmed.add0484

7. Whitcroft KL. Assessment of Human Olfaction: Investigations into Clinical Practice and Neuroanatomical Correlates of Dysfunction.

8. Seubert J, Freiherr J, Djordjevic J, Lundström JN. Statistical localization of human olfactory cortex. NeuroImage. 2013;66:333–342. doi:10.1016/j.neuroimage.2012.10.030

9. Zou L quan, van Hartevelt TJ, Kringelbach ML, Cheung EFC, Chan RCK. The neural mechanism of hedonic processing and judgment of pleasant odors: An activation likelihood estimation meta-analysis. Neuropsychology. 2016;30(8):970–979. doi:10.1037/neu0000292

10. Whitcroft KL, Mancini L, Yousry T, Hummel T, Andrews PJ. Functional septorhinoplasty alters brain structure and function: Neuroanatomical correlates of olfactory dysfunction. Front Allergy. 2023;4:1079945. doi:10.3389/falgy.2023.1079945

11. Whitcroft KL, Noltus J, Andrews P, Hummel T. Sinonasal surgery alters brain structure and function: Neuroanatomical correlates of olfactory dysfunction. J Neurosci Res. 2021;99(9):2156–2171. doi:10.1002/jnr.24897

12. Zhao K, Scherer PW, Hajiloo SA, Dalton P. Effect of anatomy on human nasal air flow and odorant transport patterns: implications for olfaction. Chem Senses. 2004;29(5):365–379. doi:10.1093/chemse/bjh033

13. Wingrove J, Makaronidis J, Prados F, et al. Aberrant olfactory network functional connectivity in people with olfactory dysfunction following COVID-19 infection: an exploratory, observational study. eClinicalMedicine. 2023;58. doi:10.1016/j.eclinm.2023.101883

14. Pendolino AL, Scarpa B, Andrews PJ. The Effectiveness of Functional Septorhinoplasty in Improving COVID-19-related Olfactory Dysfunction. Facial Plast Surg. 2025;42(1):17–27. doi:10.1055/a-2535-0153

15. Pendolino AL, Nardello E, Lund VJ, et al. Comparison between unilateral PNIF and rhinomanometry in the evaluation of nasal cycle. Rhinology. 2018;56(2):122–126. doi:10.4193/Rhin17.168

16. Whitcroft KL, Cuevas M, Haehner A, Hummel T. Patterns of olfactory impairment reflect underlying disease etiology. The Laryngoscope. 2017;127(2):291–295. doi:10.1002/lary.26229

17. Oleszkiewicz A, Schriever VA, Croy I, Hähner A, Hummel T. Updated Sniffin’ Sticks normative data based on an extended sample of 9139 subjects. Eur Arch Oto-Rhino-Laryngol Off J Eur Fed Oto-Rhino-Laryngol Soc EUFOS Affil Ger Soc Oto-Rhino-Laryngol - Head Neck Surg. 2019;276(3):719–728. doi:10.1007/s00405-018-5248-1

18. Gudziol V, Lötsch J, Hähner A, Zahnert T, Hummel T. Clinical significance of results from olfactory testing. The Laryngoscope. 2006;116(10):1858–1863. doi:10.1097/01.mlg.0000234915.51189.cb

19. Lübke K, Gottschlich M, Gerber J, Pause BM, Hummel T. No Effects of Handedness on Passive Processing of Olfactory Stimuli: An FMRI Study. Chemosens Percept. 2012;5(1):22–26. doi:10.1007/s12078-011-9115-3

20. Whitcroft KL, Mancini L, Yousry T, Hummel T, Andrews PJ. Functional septorhinoplasty alters brain structure and function: Neuroanatomical correlates of olfactory dysfunction. Front Allergy. 2023;4:1079945. doi:10.3389/falgy.2023.1079945

21. Gaviraghi M, Lupi E, Grosso E, et al. The Sense of Smell (SoS) Atlas: Its Creation and First Application to Investigate COVID-19 Related Anosmia With a Comprehensive Quantitative MRI Protocol. J Magn Reson Imaging. 2026;63(2):574–593. doi:10.1002/jmri.70128

22. Pendolino AL, Scarpa B, Andrews PJ. The Effectiveness of Functional Septorhinoplasty in Improving COVID-19-related Olfactory Dysfunction. Facial Plast Surg. 2025;42(1):17–27. doi:10.1055/a-2535-0153

23. Arnold TC, You Y, Ding M, Zuo XN, Araujo I de, Li W. Functional Connectome Analyses Reveal the Human Olfactory Network Organization. eNeuro. 2020;7(4). doi:10.1523/ENEURO.0551-19.2020

24. Torske A, Koch K, Eickhoff S, Freiherr J. Localizing the human brain response to olfactory stimulation: A meta-analytic approach. Neurosci Biobehav Rev. 2022;134:104512. doi:10.1016/j.neubiorev.2021.12.035

25. Zhang H, Chung TWH, Wong FKC, Hung IFN, Mak HKF. Changes in the Intranetwork and Internetwork Connectivity of the Default Mode Network and Olfactory Network in Patients with COVID-19 and Olfactory Dysfunction. Brain Sci. 2022;12(4):511. doi:10.3390/brainsci12040511

26. Han P, Croy I, Raue C, et al. Neural processing of odor-associated words: an fMRI study in patients with acquired olfactory loss. Brain Imaging Behav. 2020;14(4):1164–1174. doi:10.1007/s11682-019-00062-2

27. Abdul Manan H, de Jesus R, Thaploo D, Hummel T. Mapping the Olfactory Brain: A Systematic Review of Structural and Functional Magnetic Resonance Imaging Changes Following COVID-19 Smell Loss. Brain Sci. 2025;15(7):690. doi:10.3390/brainsci15070690

28. Gaviraghi M, Lupi E, Grosso E, et al. The Sense of Smell (SoS) Atlas: Its Creation and First Application to Investigate COVID-19 Related Anosmia With a Comprehensive Quantitative MRI Protocol. J Magn Reson Imaging JMRI. 2026;63(2):574–593. doi:10.1002/jmri.70128

29. Eek T, Lundin F, Larsson M, Hamilton P, Georgiopoulos C. Neural suppression in odor recognition memory. Chem Senses. 2023;48:bjad001. doi:10.1093/chemse/bjad001

30. Ruser P, Koeppel CJ, Kitzler HH, Hummel T, Croy I. Individual odor hedonic perception is coded in temporal joint network activity. NeuroImage. 2021;229:117782. doi:10.1016/j.neuroimage.2021.117782

